# NMDAR subunit GluN1 dynamics in urinary astrocyte-derived extracellular vesicles in anti-NMDAR encephalitis

**DOI:** 10.64898/2026.03.30.26349276

**Authors:** Junhua Mei, Mian-mian Chen, Qing Yang, Shu-xian Xu, Chao Wang, Honggang Lyu, Qian Gong, Zhongchun Liu, Edward Bullmore, Mary-Ellen Lynall, Xin-hui Xie

**Affiliations:** Department of Neurology, Wuhan First Hospital, Wuhan, Hubei, China; Department of Psychiatry, Renmin Hospital of Wuhan University, Wuhan, China; State Key Laboratory of Metabolism and Regulation in Complex Organisms, Wuhan University, Wuhan, China; Department of Psychiatry, School of Clinical Medicine, University of Cambridge, Cambridge, UK; School of Academic Psychiatry, Institute of Psychiatry, Psychology & Neuroscience, King’s College London, London, UK; Department of Psychiatry, University of Oxford, Oxford, UK; Sanger Institute, Wellcome Genome Campus, Hinxton, UK

**Keywords:** exosomes, proteomics, urine, auto-antibody

## Abstract

**Background and Objectives:** Longitudinal measurement *in vivo* of molecular changes in the central nervous system (CNS) is important to investigate and guide treatment of rapidly changing neurological diseases. Here, we show how a novel protocol for urinary astrocyte-derived extracellular vesicles (uADEVs) can be used non-invasively to track the dynamic trajectory of the N-methyl-D-aspartate receptor (NMDAR) subunit GluN1 in a patient with anti-NMDAR encephalitis.

**Methods:** We collected 18 morning urine samples from the patient over 34 days of hospitalisation, as well as 15 samples from a healthy control. We isolated uADEVs, measured GluN1 concentrations, and used a wavelet transform to analyse the time-frequency dynamics of the GluN1 signal.

**Results:** Compared with the control, GluN1 levels in the patient’s uADEVs were higher on average and more variable over time. Wavelet analysis resolved (i) a low-frequency trend that decreased over time, matching the decrease of GluN1 in cerebrospinal fluid associated with clinical recovery; and (ii) a higher frequency oscillation, with GluN1 levels peaking about 48-72 hours after each of 4 cycles of methotrexate infusion.

**Conclusion:** This case study demonstrates the feasibility of urinary ADEV sampling to measure brain biomarker dynamics linked to the clinical course and treatment of auto-immune encephalitis.

## Introduction

Anti-NMDA receptor encephalitis is thought to arise from antibody-induced NMDAR internalization and consequent reductions in receptor density – a mechanistic understanding primarily based on *in vitro* and animal studies (1,2). Direct human evidence was limited to postmortem tissue staining until a recent positron emission tomography (PET) study (3) provided the first such evidence *in vivo*. Compared with healthy controls, patients with serum antibody-positive anti-NMDAR encephalitis showed reduced NMDAR density, whereas serum antibody-negative patients did not. However, PET has three major limitations: scans are necessarily infrequent making rapid changes during the acute stage difficult to track; the signal cannot be assigned to a specific cell type; and brain scanning is poorly tolerated in acute encephalitis.

Assays based on brain-derived extracellular vesicles (BDEVs) offer an alternative solution (4). These nanosized particles encapsulate proteins, lipids, and nucleic acids from intracellular cytoplasm and can cross the blood-brain barrier bidirectionally, appearing in blood. They can therefore serve as peripherally accessible proxies for the molecular status of specific brain cells. For example, in Alzheimer’s (5) and Parkinson’s disease (6), disease-specific biomarkers measured in plasma BDEVs strongly correlate with their measurements in CNS-adjacent CSF.

We have recently shown that BDEVs are also measurable in urine (7). Here, we report for the first time how our innovative protocol for urinary astrocyte-derived extracellular vesicle (uADEV) isolation can be used to measure the dynamics of brain glutamatergic receptor protein in a patient over the course of treatment for anti-NMDAR encephalitis. This single case had a clear diagnosis, a dynamic disease course, and a well-regulated treatment plan, which provided a robust basis for interpreting brain molecular signals measured in uADEVs isolated from urine sampled repeatedly over time.

### Case Report

A 30-35-year-old female was admitted to hospital in Wuhan, China with a 12-day history of psychiatric symptoms and a 3-day history of involuntary limb movements and altered consciousness. On admission (day 0), she was diagnosed with anti-NMDAR encephalitis, with a CSF anti-NMDAR antibody titre of 1:100. As shown in **Figure 1**, she subsequently received four intravenous infusions of human gamma-globulin (the first four red dotted vertical lines), four cycles of intrathecal methotrexate (the four solid yellow vertical lines), four cycles of intravenous rituximab (the four blue dashed vertical lines), and a tapering dose of methylprednisolone (the green horizontal line at the top of the subfigures). The patient regained full consciousness on day 19 and was discharged on day 34, when her CSF antibody titre was undetectable. She returned to the hospital for a follow-up visit three months after discharge.

**Figure 1.**
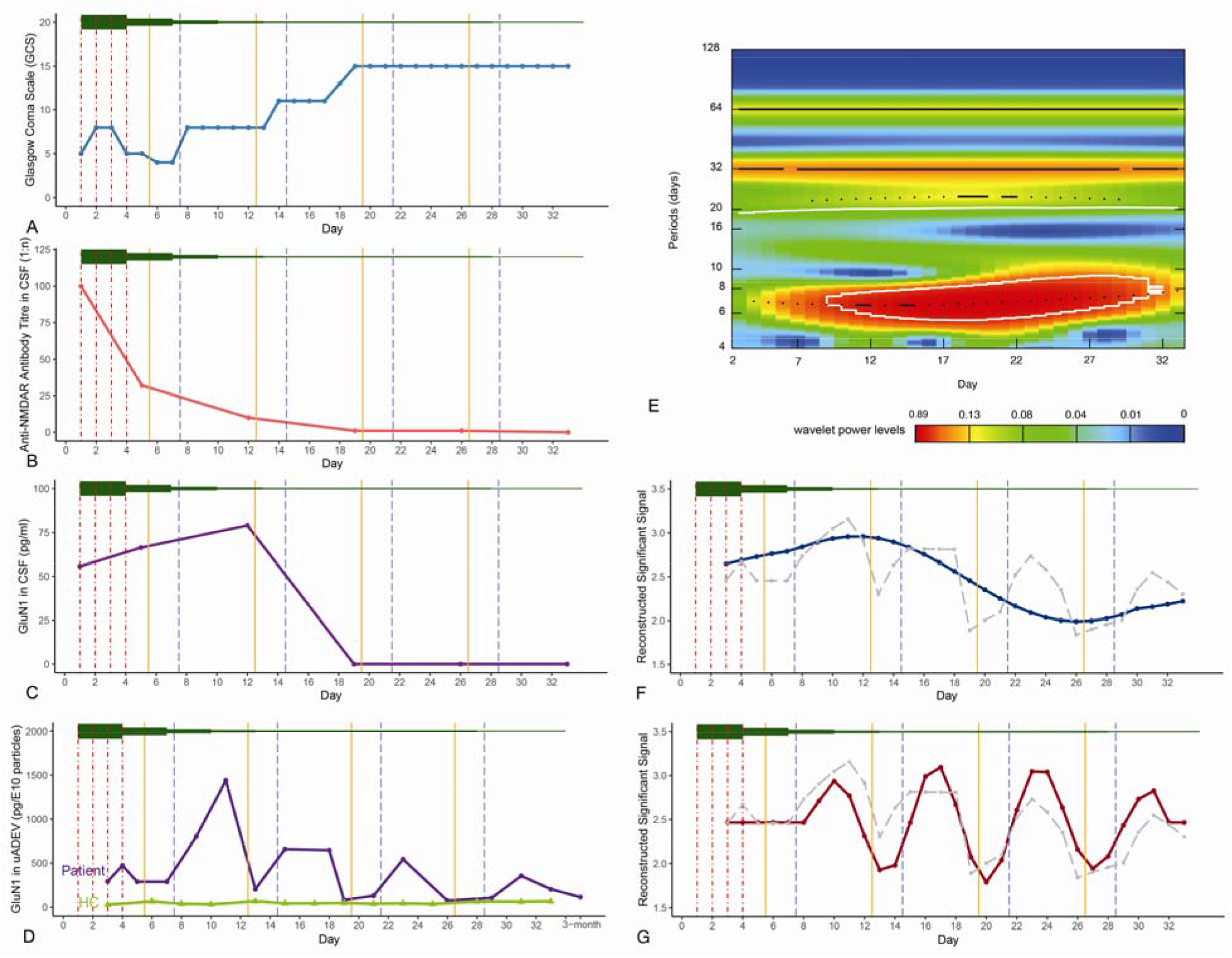
Clinical timeline and dynamics of GluN1 measured in urinary astrocyte-derived extracellular vesicles (uADEVs) from a single case of anti-NMDAR encephalitis and a healthy control. On admission (day 0), the patient was diagnosed with anti-NMDAR encephalitis and received four intravenous infusions of human gamma-globulin (first four red dotted vertical lines), four cycles of intrathecal methotrexate (MTX; four solid yellow vertical lines), four cycles of intravenous rituximab (four blue dashed vertical lines), and a tapering dose of methylprednisolone (green horizontal line at the top of the subfigures). These treatment indicators are the same in all subfigures. (**A**) Glasgow Coma Scale (GCS) scores (y-axis) were repeatedly measured over the course of in-patient treatment (days 0-33, x-axis), showing sustained recovery from about day 20. There was clear evidence of reduced titres of anti-NMDAR antibody (NMDAR-Ab) (**B**), and reduced concentrations of GluN1 (**C**), in six cerebrospinal fluid (CSF) samples collected over the course of treatment. We concurrently measured the concentration of GluN1 in ADEVs (pg/per 10^10^ particles by nanoparticle tracking analysis (NTA); y-axis) isolated from 18 urine samples from the encephalitis case (**D**, purple line) and compared this dynamic profile to repeated GluN1 measurements from 15 urinary ADEVs in the healthy control subject (**D**, light green line). Time-frequency decomposition by wavelet analysis of the log_10_(GluN1) time series was represented by a scalogram of variance as a function of frequency (y-axis) and time (x-axis). We interpolated values to a daily scale by averaging adjacent data points because intervals varied between 1-2 days (**E**). This analysis highlighted two dynamic processes: a low frequency trend to lower levels of GluN1 over the course of treatment (**F**), and a high frequency oscillation closely coupled to the timing of methotrexate infusions (**G**). The gray dashed lines in (**F**) and (**G**) represent the log_10_(ADEV GluN1) time series.

The molecular target of autoantibodies generated during anti-NMDAR encephalitis is typically the GluN1 subunit of the glutamatergic NMDA receptor (2). As methods for isolating ADEVs from urine are more mature than methods for urinary neuron-derived EVs, and because astrocytes are known to express NMDA receptors (8), we predicted that urinary ADEVs could provide relevant information on the changing status of GluN1 over the course of this clinical case. During the 34 days of hospitalization, we prospectively collected morning urine samples approximately every two days and isolated ADEVs from these 18 samples. Detailed isolation procedures are described in our previous methodological study (7). We also recruited a healthy female volunteer aged 30–35 years, collected 15 urine samples at two-day intervals, and isolated uADEVs using the same protocol. We then measured GluN1 concentrations in lysed ADEVs from the 18 patient samples, 15 control samples, and seven CSF samples from the patient, using an enzyme-linked immunosorbent assay (CUSABIO, Catalog# CSB-EL009911HU). To normalize the GluN1 concentrations, we measured the particle number of each uADEV sample using nanoparticle tracking analysis. **Figure 1** shows the complete treatment course and changes in the various biological measurements.

We hypothesized that GluN1 levels would change dynamically in relation to clinical treatment and recovery. It was evident by inspection of the GluN1 protein concentrations repeatedly measured in uADEVs (**Figure 1D**) that GluN1 levels were on average higher and more variable over time in the case of encephalitis compared to the healthy control. To resolve the dynamics more precisely, we used the Morlet wavelet transform for time-frequency analysis of the GluN1 time series (**Figure 1E**). The wavelet transform captures how frequency content evolves over time, providing a richer picture of the signal’s dynamics than time-averaged approaches like Fourier analysis. Two clear dynamics emerged: (i) a low frequency curvilinear trend from higher GluN1 concentrations in the first half of the inpatient treatment period (0-15 days) to lower levels in the second half (16-33 days) (**Figure 1F**), mirroring the step-down from high CSF concentrations of GluN1 over the same period (**Figure 1C**); and (ii) a high frequency oscillation which was coupled in time to the schedule of methotrexate infusions, with an upward inflection in GluN1 concentrations immediately post infusion (**Figure 1G**), reaching a peak about 48h after the start of each infusion cycle.

## Discussion

It may seem counter-intuitive that increased levels of GluN1 should be associated with both increased clinical severity of this auto-immune disease and with response to its effective treatment. However, these observations can be hypothetically reconciled in terms of a simple model of endosomal, lysosomal and exosomal processing of NMDA receptors that have been compromised by anti-NMDAR auto-antibody binding or linkage.

Membrane receptors are internalised by invagination (1) of the cell surface to form membrane-bound intracellular vesicles or endosomes, which then pass through the multivesicular body (MVB) pathway, leading either to degradation by lysosomes or to extracellular release as exosomes (9). Autoantibody binding upregulates this process nearly 8-fold (10). Early in the treatment period, when the titre of anti-NMDA auto-antibodies was high (**Figure 1B**), it is therefore predictable that there would be an increased number of endosomes being processed through the MVB pathway to be released as GluN1-enriched exosomes into the extracellular space of the brain and thence into the blood circulation and ultimately the urine.

However, the number of ADEVs measurable in the circulation does not depend solely on the rate of NMDAR internalization to form endosomes but also on the proportion of endosomes that are switched by the MVB pathway to lysosomal destruction versus exosomal release. The switch to exosomal release can be promoted by p53 (11), a regulatory transcription factor protein that has been implicated in tumour suppression and many cell cycle processes. Methotrexate can boost p53 protein levels, peaking ∼24-48h post-dose (12), causing increased exosome production over the same period (11). Thus, the repeated cycle of increased GluN1 following methotrexate infusion can be explained by the drug’s known effects on p53 causing a greater proportion of GluN1-enriched endosomes to be released as exosomes, and therefore measurable in urine as ADEVs. In support of this model, it is notable that the maximum methotrexate-driven level of GluN1 was typically measured in urinary ADEVs about 48-72h after dosing, i.e., it was delayed by 12-24h relative to the known methotrexate-driven peak in p53 (11,12). This time lag is approximately compatible with the time taken for brain-derived EVs to enter the bloodstream and then to be excreted in the urine.

## Conclusion

This logically coherent pattern of results from clinical investigation of a single case will clearly require replication in larger samples of patients with anti-NMDAR encephalitis, and deeper mechanistic understanding of exosome pathophysiology from future cellular and experimental medicine studies. However, this first case report (N=1) suggests that urinary ADEV sampling could provide a clinically acceptable and informative proxy for dynamic fluxes in astrocyte NMDAR signaling, exemplifying the diagnostic potential of this novel technique for reading “a message in a bottle” from the brain.

## Data Availability

The data that support the findings of this study are available from the corresponding author upon reasonable request.

## Declarations

Ethics approval and consent to participate: This study was conducted at Wuhan First Hospital, Wuhan, People’s Republic of China, in compliance with the Declaration of Helsinki (revised edition, 2013). The patient and her legal guardian as well as the healthy control provided informed consents and were free to withdraw their consent at any time for any reason.

## Code availability

The source code for this study is available at GitHub: https://github.com/xxh87/uADEV_anti_NMDAR_Encephalitis_Case.

## References

1. Dalmau J, Graus F (2018): Antibody-Mediated Encephalitis. N Engl J Med 378: 840–851.

2. Dalmau J, Armangué T, Planagumà J, Radosevic M, Mannara F, Leypoldt F, et al. (2019): An update on anti-NMDA receptor encephalitis for neurologists and psychiatrists: mechanisms and models. Lancet Neurol 18: 1045–1057.

3. Galovic M, Al-Diwani A, Vivekananda U, Walker M C, Irani S R, Koepp, M J, et al (2023). In Vivo N-Methyl-d-Aspartate Receptor (NMDAR) Density as Assessed Using Positron Emission Tomography During Recovery From NMDAR-Antibody Encephalitis. JAMA Neurol:80, 211–213.

4. Manolopoulos A, Yao PJ, Kapogiannis D (2025): Extracellular vesicles: translational research and applications in neurology. Nat Rev Neurol 21: 265–282.

5. Jia L, Qiu Q, Zhang H, Chu L, Du Y, Zhang J, et al. (2019): Concordance between the assessment of Aβ42, T-tau, and P-T181-tau in peripheral blood neuronal-derived exosomes and cerebrospinal fluid. Alzheimer’s & Dementia 15: 1071–1080.

6. Jiang C, Hopfner F, Berg D, Hu MT, Pilotto A, Borroni B, et al. (2021): Validation of αLJSynuclein in L1CAMLJImmunocaptured Exosomes as a Biomarker for the Stratification of Parkinsonian Syndromes. Movement Disord mds.28591.

7. Xie X-H, Chen M-M, Xu S-X, Mei J, Yang Q, Wang C, et al. (2025): Isolating Astrocyte-Derived Extracellular Vesicles From Urine. Int J Nanomed 20: 2475–2484.

8. Skowrońska K, Obara-Michlewska M, Zielińska M, Albrecht J (2019): NMDA Receptors in Astrocytes: In Search for Roles in Neurotransmission and Astrocytic Homeostasis. Int J Mol Sci 20: 309.

9. Scita G, Di Fiore PP (2010): The endocytic matrix. Nature 463: 464–473.

10. Moscato EH, Peng X, Jain A, Parsons TD, Dalmau J, Balice-Gordon RJ (2014): Acute mechanisms underlying antibody effects in anti-N-methyl-D-aspartate receptor encephalitis. Ann Neurol 76: 108–119.

11. Yu X, Harris SL, Levine AJ (2006): The regulation of exosome secretion: a novel function of the p53 protein. Cancer Res 66: 4795–4801.

12. Huang W-Y, Yang P-M, Chang Y-F, Marquez VE, Chen C-C (2011): Methotrexate induces apoptosis through p53/p21-dependent pathway and increases E-cadherin expression through downregulation of HDAC/EZH2. Biochem Pharmacol 81: 510–517.

